# Life course traumas, phenotypic aging, and cardiovascular disease: retrospective analysis of 104,939 UKB participants

**DOI:** 10.1101/2021.11.24.21266842

**Authors:** Xingqi Cao, Jingyun Zhang, Chao Ma, Xueqin Li, Chia-Ling Kuo, Morgan E. Levine, Guoqing Hu, Heather Allore, Xi Chen, Xifeng Wu, Zuyun Liu

**Affiliations:** Center for Clinical Big Data and Analytics, Second Affiliated Hospital and Department of Big Data in Health Science, School of Public Health, Zhejiang University School of Medicine, Hangzhou 310058, Zhejiang, China; School of Economics and Management, Southeast University, Nanjing 211189, Jiangsu, China; Department of Community Medicine and Health Care, Connecticut Convergence Institute for Translation in Regenerative Engineering, Institute for Systems Genomics, University of Connecticut Health, Farmington, CT 06030, USA; Department of Pathology, Yale School of Medicine, New Haven, CT 06520, USA; Department of Epidemiology and Health Statistics, Xiangya School of Public Health, Central South University, Changsha 410083, Hunan, China; National Clinical Research Center for Geriatric Disorders, Xiangya Hospital, Central South University, Changsha 410008, Hunan, China; Department of Internal Medicine, Yale School of Medicine, New Haven, CT 06520, USA; Department of Health Policy and Management, Yale School of Public Health, New Haven, CT 06520, USA; Department of Economics, Yale University, New Haven, CT 06520, USA

**Keywords:** Traumas, Life course, Phenotypic aging, Cardiovascular disease

## Abstract

**Background:** While childhood and adulthood traumatic experiences have been linked to subsequent cardiovascular disease (CVD), the relationship between life course traumas and CVD and the underpinning pathways are poorly understood. This study aimed to: (1) examine the associations of childhood, adulthood, and lifetime traumas with CVD; (2) examine the associations between diverse life course traumatic profiles and CVD; and (3) examine the extent to which Phenotypic Age (PhenoAge), a well-developed phenotypic aging measure, mediates these associations.

**Methods:** We included 104,939 participants from the UK Biobank who completed the 2016 online mental health questionnaire. CVD outcomes including ischemic heart disease, myocardial infarction, and stroke were ascertained. Childhood, adulthood, and lifetime traumas were categorized into three subgroups (mild, moderate, and severe), respectively. Four life course traumatic profiles were defined as *non-severe traumas across life course, non-severe childhood and severe adulthood traumas, severe childhood and non-severe adulthood traumas, and severe traumas across life course* based on both childhood and adulthood traumas. PhenoAge was measured using an equation previously developed. Multivariable logistic models and formal mediation analyses were performed.

**Results:** Of 104,939 participants, 7,398 (7.0%) were diagnosed with CVD. Subgroups of childhood, adulthood, and lifetime traumas were associated with CVD, respectively. Furthermore, life course traumatic profiles were significantly associated with CVD. For instance, compared with subgroups experiencing *non-severe traumas across life course*, those who experienced *non-severe childhood and severe adulthood traumas, severe childhood and non-severe adulthood traumas, and severe traumas across life course* had higher odd of CVD, with odds ratios of 1.07 (95% confidence interval [CI]: 1.00, 1.15), 1.17 (95% CI: 1.09, 1.25), and 1.33 (95% CI: 1.24, 1.43), respectively. Formal mediation analyses suggested that PhenoAge partially mediated the above associations. For instance, PhenoAge mediated 5.8% of increased CVD events in subgroups who experienced severe childhood traumas, relative to those experiencing mild childhood traumas.

**Conclusions:** Childhood, adulthood, and lifetime traumas, as well as diverse life course traumatic profiles, were associated with CVD. Furthermore, phenotypic aging partially mediated these associations. These findings suggest a potential pathway from life course traumas to CVD through phenotypic aging, and underscore the importance of policy programs targeting traumatic events over the life course in ameliorating inequalities in cardiovascular health.

## Introduction

Cardiovascular disease (CVD) including ischemic heart disease (IHD) and stroke is the leading cause of mortality, accounting for about 27% of global deaths [1]. Emerging evidence have consistently recognized early life traumas (e.g., physical abuse, emotional neglect) as potent contributors to CVD [2-5], while with limited emphasis given to life course (both childhood and adulthood) traumatic experiences [6]. A life course approach address how life course exposures affect health inequalities using critical periods [7], accumulative risk [7], and chains of risk models [8]. This approach may help identify subpopulations with increased health risks due to unique life course traumatic profiles, e.g., those who experienced severe childhood and non-severe adulthood traumas, and this, however, has not been investigated.

How life course exposures “get under the skin” and result in the onset of chronic diseases also remains unclear. It is highly possible that traumas lead to physiological dysregulation [9, 10], manifesting as vulnerability to disease, which in turn increases the risk of CVD. Aging is one of the major risk factors for most chronic diseases. It is intuitive to establish a pathway, life course exposures → aging → chronic disease, in line with the Geroscience paradigm [11]. In fact, it has been reported that traumas accelerate aging at both cellular (e.g., shorten telomere length) [12] and phenotypic levels (e.g., Phenotypic Age Acceleration [PhenoAgeAccel]) [13]. Though phenotypic aging predicts chronic diseases [14, 15], no studies have rigorously evaluated the extent to which phenotypic aging mediates the association between life course traumas and CVD.

In this study, based on data from a large sample of middle-aged and older adults participating in the UK Biobank (UKB) study, we grouped participants in two ways: (1) defining three subgroups (mild, moderate, and severe) based on separate assessments on childhood, adulthood, and lifetime traumas, respectively; (2) defining four life course traumatic profiles as *non-severe traumas across life course, non-severe childhood and severe adulthood traumas, severe childhood and non-severe adulthood traumas, and severe traumas across life course* based on both childhood and adulthood traumas. On account of the evidence that traumas may accelerate aging [12, 13] and the fact that aging is a major risk factor for CVD, we hypothesized that life course traumas may affect cardiovascular health through aging. We first examined the associations of childhood, adulthood, and lifetime traumas (i.e., three subgroups, respectively) with CVD. Next, we examined the association of four life course traumatic profiles with CVD. Finally, with Phenotypic Age (PhenoAge), a well-developed phenotypic aging measure that captures morbidity and mortality risk [14, 15], we performed formal mediation analyses to examine the extent to which PhenoAge mediates these associations above. The findings would deepen the understanding of the complex associations between life course traumas, phenotypic aging, and cardiovascular health, and thus provide clues for implementing policy programs.

## Materials and Methods

### Study participants

UKB study is an ongoing, population-based cohort study of about half a million adults aged 40-69 years in the UK, beginning in 2006 [16]. UKB was approved by the North West Multi-Centre Research Ethics Committee, the National Information Governance Board for Health & Social Care in England and Wales and the Community Health Index Advisory Group in Scotland. Informed consent was obtained from all participants. In 2016, about two-thirds of the participants were invited to complete an online mental health questionnaire due to the availability of their email addresses [17]. Less than half of them responded to this questionnaire (N=156,749). Participants with incomplete data on life course traumas (N=10,891), clinical biomarkers (N=24,337), or covariates (N=16,582) were excluded, resulting in 104,939 participants included in our analyses.

### Assessment of childhood, adulthood, and lifetime traumas

Traumas involving childhood, adulthood, and lifetime were assessed through the 2016 online mental health questionnaire survey (**Figure 1**).

**Figure 1.**
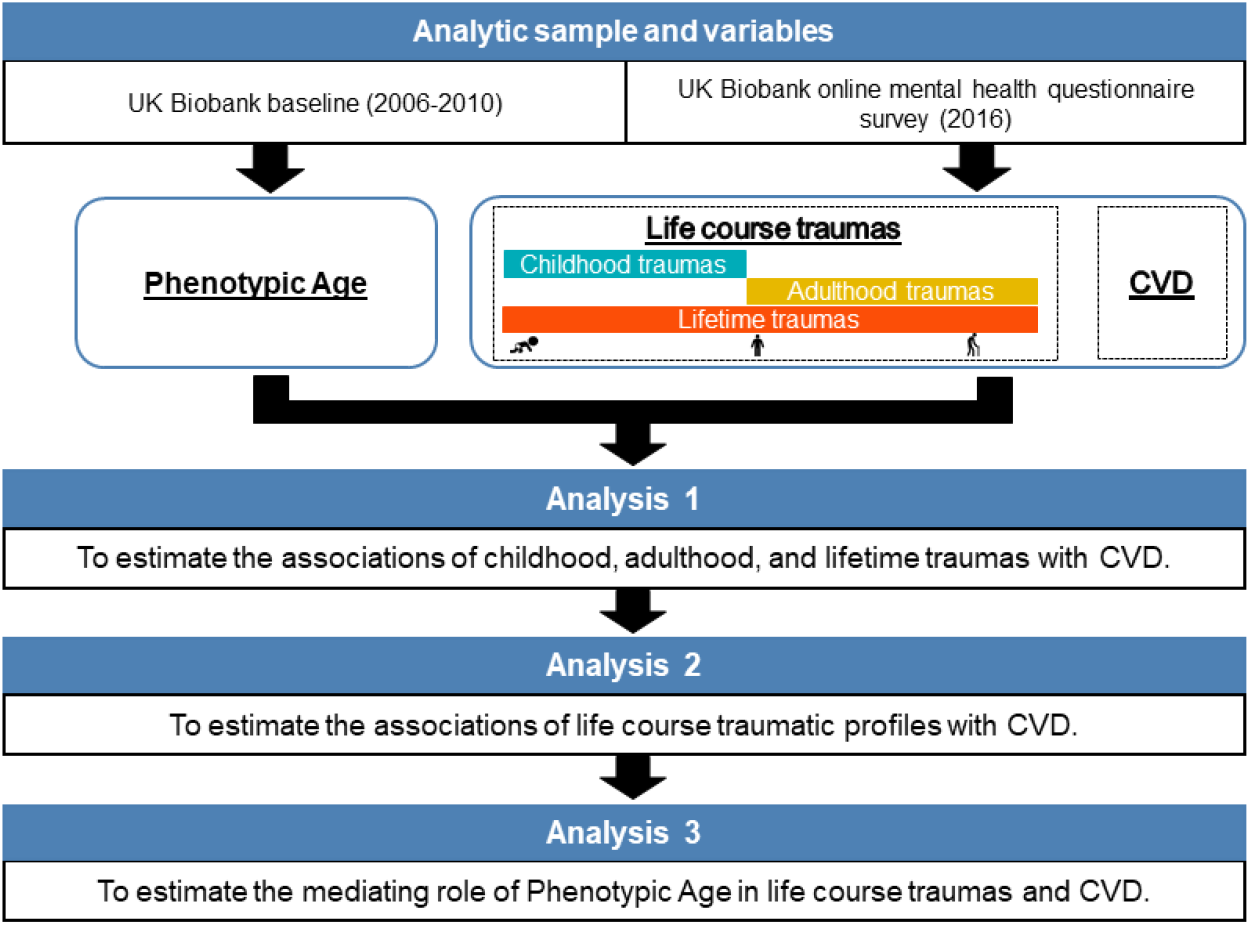
Roadmap for evaluating the complex associations between life course traumas, phenotypic aging, and CVD. The roadmap depicts the analytic sample, variables and our analytical procedures. CVD, cardiovascular disease.

Childhood traumas were assessed using the childhood trauma screener (CTS), which covered sexual, emotional and physical abuse, and emotional and physical neglect when growing up [18] (**Table S1**). The response for each item was set on a five-point Likert scale: “never true”, “rarely true”, “sometimes true”, “often”, and “very often true”, with a score of 0-4. The total score ranged from 0 to 20.

Adulthood traumas, which were those that occurred after 16 years of age, were assessed using six items devised by the UKB study team. Experiences of intimate partner violence, relationship and financial insecurity, and sexual and physical abuse during adulthood were covered (**Table S1**). To be comparable to the CTS, adulthood trauma items using the same five-point Likert scale with a score of 0-4. The total score ranged from 0 to 20.

Lifetime traumas were assessed by a short checklist as defined by the Diagnostic and Statistical Manual of Mental Disorders [19, 20]. Participants were asked whether they had ever: “experienced serious accident”, “involved in combat or exposed to war-zone”, “witnessed sudden violent death”, “experienced physical abuse”, “experienced sexual abuse”, and “diagnosed with life-threatening illness” in life (**Table S1**). The response for recent lifetime traumas was categorized into “never”, “yes, but not in the last 12 months”, and “yes, within the last 12 months”, with a score of 0-2. Thus, the total score ranges from 0 to 12.

To assess the accumulative traumatic experiences across the life course, we summarized childhood, adulthood, and lifetime traumas to obtain an “all traumas” variable, ranging from 0 to 52.

Since traumas variables were correlated (**Figure 2**), we used principal component analysis (PCA) to integrate them to obtain a composite measure. We ran PCA for childhood traumas, and then used the first principal component (PC1) to define three subgroups (mild, moderate, and severe), corresponding to the lowest, middle, and highest tertile of PC1. We defined subgroups for adulthood and lifetime traumas similarly. Furthermore, based on the chains of risk model, we defined four life course traumatic profiles: (1) *non-severe traumas across life course* (proportion of participants, 52.5%); (2) *non-severe childhood and severe adulthood traumas* (18.0%); (3) *severe childhood and non-severe adulthood traumas* (15.2%); (4) *severe traumas across life course* (14.4%), based on subgroups of both childhood and adulthood traumas (without information on lifetime traumas, since the latter reflects confusing timeframe; **Table S2**).

**Figure 2.**
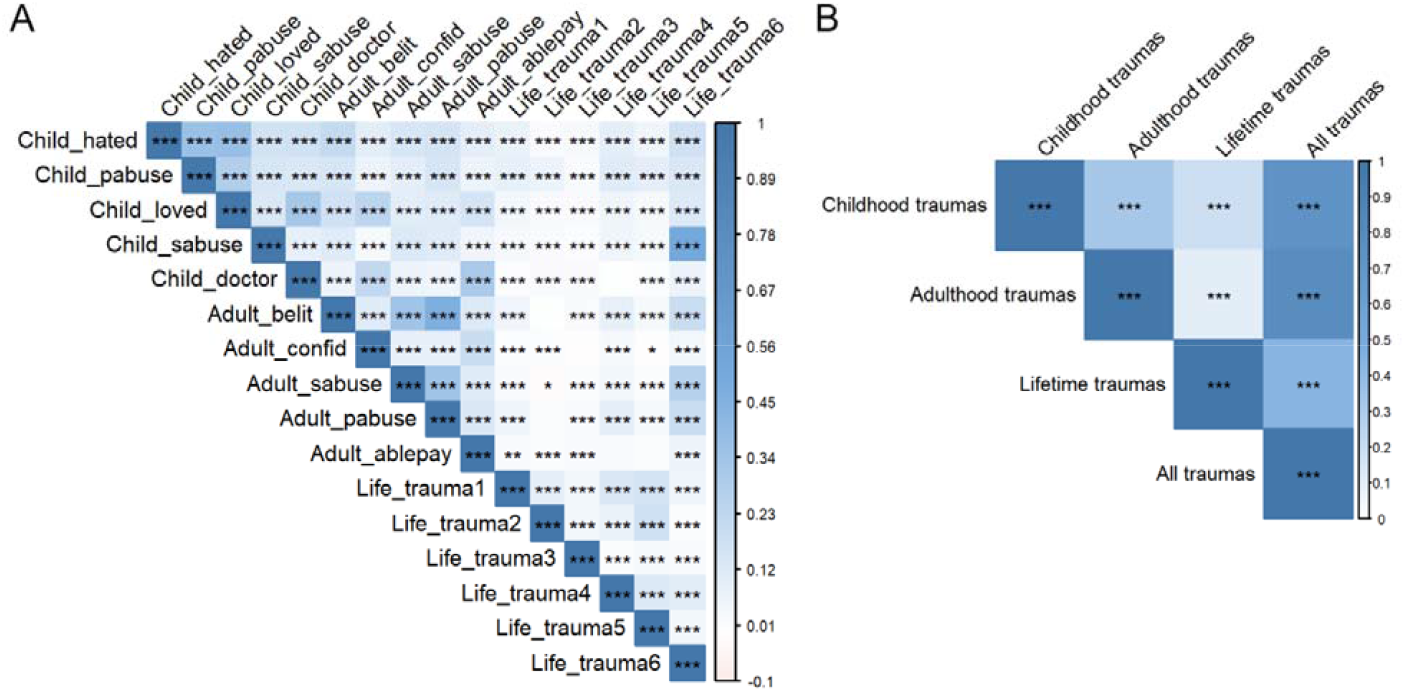
Correlations of life course traumas items and summary scores assessed in the UKB. (A) Each cell shows the spearman correlation (and P value) between items for life course traumas. (B) Each cell shows the spearman correlation (and P value) between summary scores for life course traumas. The closer to “dark blue”, the higher correlation coefficient among these two items (or summary scores). *, P<0.05; **, P <0.01, ***, P<0.001. Details on these items can be found in Table S1.

### Assessment of phenotypic aging

We calculated PhenoAge according to procedures we developed in previous studies [15]. PhenoAge has been applied to UKB participants in our recent work [21]. In brief, nine biomarkers and chronological age collected at baseline were used to calculate PhenoAge (in years), with the equation as follows:

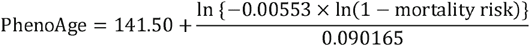

where

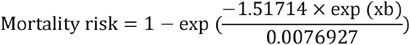

and

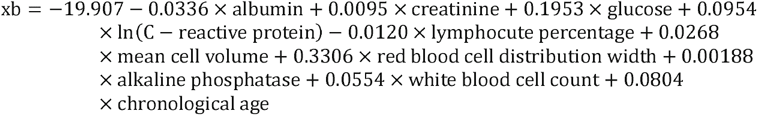

### CVD

The diagnosis of CVD, including IHD, myocardial infarction (MI), and stroke, was defined at the time of attending the 2016 online mental health questionnaire survey. We combined information of the International Statistical Classification of Diseases and Related Health Problems 9th (ICD-9) (IHD: 410, 411, 412, 413, 414; MI: 410, 411, 412, 429.79; Stroke: 430, 431, 432, 433, 434, 435, 436, 437, and 438) and 10th (ICD-10) (IHD: I20, I21, I22, I23, I24, I25; MI: I21, I22, I23, I24.1, I25.2; Stroke: I60, I61, I63, and I64) revisions (obtained through linked hospital admissions data), and self-reported outcomes [22], which was in line with algorithmic definitions of UKB.

### Other covariates

Other covariates including chronological age, sex, ethnicity, educational level, occupation, Townsend deprivation index (TDI), smoking status, alcohol consumption, regular exercise, maternal smoking around birth, and family history of CVD were obtained through a questionnaire at baseline. Ethnicity was defined as White, Mixed, Asian, Black, Chinese, and others. Educational level was defined as high (i.e., college or university degree), intermediate (i.e., A/AS levels or equivalent, O levels/ General Certificate of Secondary Education levels or equivalent), and low (none of the above). The occupation was defined as working, retired, and others (including unemployed, student, volunteer, and so on). TDI was a composite score based on information of unemployment, overcrowding, home ownership, and car ownership [23]. A higher score of TDI indicates a lower level of socioeconomic status [23]. Smoking status was defined as non-smoker, former smoker, and current smoker. Alcohol consumption was defined as never or occasional, 1-3 times/month, 1-4 times/week, and daily. Regular exercise was defined as yes (75 minutes of vigorous activity or 150 minutes of moderate activity or an equivalent combination per week), and no [24]. We used maternal smoking around birth (yes vs no) to reflect childhood socioeconomic status [25]. Family history of CVD (yes or no) was also considered. Additionally, body mass index (BMI) as weight (kg)/height (m)^2^ was also obtained at baseline.

### Statistical analyses

Socio-demographic characteristics were described with mean ± standard deviation (SD) and number (percentage) for continuous and categorical variables, respectively.

As shown in **Figure 1**, we first estimated the associations between life course traumas and CVD. We used the Shapley Value Decomposition method to estimate the overall and relative contributions of all life course traumas to CVD [26, 27]. Then, multivariable logistic regression models were used to estimate the associations of childhood, adulthood, and lifetime traumas subgroups with CVD, and adjusted odds ratios (aORs) and corresponding 95% confidence intervals (CIs) were documented from three models. Model 1 adjusted for chronological age and sex. Model 2 additionally adjusted for ethnicity, educational level, occupation, TDI, BMI, smoking status, alcohol consumption, regular exercise, maternal smoking around birth, and family history of CVD. Model 3 included all covariates of Model 2 and PhenoAge. Subsequently, we used similar models to estimate the associations between four life course traumatic profiles and CVD.

We performed three steps to examine the mediating role of PhenoAge in the associations of traumas with CVD. First, multivariable logistic regression models were used to examine the associations of PhenoAge with CVD in the full sample: model 1 adjusted for chronological age and sex; model 2 further adjusted for ethnicity, educational level, occupation, TDI, BMI, smoking status, alcohol consumption, regular exercise, maternal smoking around birth, and family history of CVD. These associations stratified by subgroups of childhood traumas, adulthood traumas, lifetime traumas, and all traumas were also estimated. Second, linear regression models were used to estimate the associations of childhood, adulthood, and lifetime traumas with PhenoAge. The aforementioned two models were used. Third, to formally estimate the extent to which PhenoAge mediated these associations, we assumed that life course traumas occurred before the baseline, while CVD occurred after the baseline (see sensitivity analysis below). The estimate was performed using the R package “mediation” [28] with 1000 simulations and the mediation proportions and corresponding 95% CIs were documented.

We performed several sensitivity analyses. First, we compared the socio-demographic characteristics between included participants and those excluded from this study in UKB. Second, we estimated the associations of the single traumatic item and summary score with CVD. Third, we focused on the original nine profiles of life course traumas based on three subgroups of childhood and adulthood traumas (**Table S3**), and estimated their associations with CVD and PhenoAge. Fourth, we excluded self-reported CVD and re-estimated the associations of life course traumas with CVD. Fifth, to strictly follow the temporality of the analytic variables (life course traumas, PhenoAge, and CVD), we only included childhood traumas and CVD events ascertained after the baseline in the mediation analysis. Cox proportional hazard regression models were performed. Finally, childhood, adulthood, lifetime, and all traumas were categorized into three subgroups according to the tertile of their corresponding summary scores (not PC1), and main analyses were repeated.

All analyses were performed using SAS version 9.4 (SAS Institute, Cary, NC), R version 3.6.3 (2020-02-29), and STATA version 14.0 software (Stata Corporation, College Station, TX). A two-sided P value <0.05 was considered to be statistically significant.

## Results

### Participants’ socio-demographic characteristics

The mean chronological age of the included participants was 56.2 (SD=7.7) years. About 55.5% were female (58,292/104,939), and the majority were White (97.2%). Almost half of the study participants had a high educational level. Only 7.0% of the study participants were current smokers, while 23.5% consumed alcohol daily. For exercise, 57.2% reported doing regular exercise. Additionally, 57.4% reported having a family history of CVD. Of 104,939 participants, 7,398 (7.0%) had a diagnosis of CVD by 2016. Detailed socio-demographic characteristics of the total participants and by CVD are presented in **Table 1**.

**Table 1.** Socio-demographic characteristics of the study participants (N=104,939).

| **Characteristics** | **Total**<br>**(N=104,939)** | **Without CVD**<br>**(N=97,541)** | **With CVD**<br>**(N=7,398)** |
| --- | --- | --- | --- |
| Chronological age, years, mean ± SD | 56.2±7.7 | 55.8±7.7 | 60.6±6.3 |
| PhenoAge, years, mean ± SD | 51.7±9.3 | 51.3±9.2 | 57.9±8.8 |
| Sex |  |  |  |
| Female, n (%) | 58,292 (55.5) | 55,851 (57.3) | 2,441 (33.0) |
| Ethnicity |  |  |  |
| White, n (%) | 102,045 (97.2) | 94,828 (97.2) | 7,217 (97.6) |
| Mixed background, n (%) | 499 (0.5) | 476 (0.5) | 23 (0.3) |
| Asian, n (%) | 870 (0.8) | 791 (0.8) | 79 (1.1) |
| Black, n (%) | 711 (0.7) | 679 (0.7) | 32 (0.4) |
| Chinese, n (%) | 237 (0.2) | 232 (0.2) | 5 (0.1) |
| Others, n (%) | 577 (0.5) | 535 (0.5) | 42 (0.6) |
| Educational level |  |  |  |
| High, n (%) | 50,114 (47.8) | 47,230 (48.4) | 2,884 (39.0) |
| Intermediate, n (%) | 34,087 (32.5) | 31,732 (32.5) | 2,355 (31.8) |
| Low, n (%) | 20,738 (19.8) | 18,579 (19.0) | 2,159 (29.2) |
| Occupation |  |  |  |
| Working, n (%) | 68,079 (64.9) | 64,648 (66.3) | 3,431 (46.4) |
| Retired, n (%) | 30,166 (28.7) | 26,735 (27.4) | 3,431 (46.4) |
| Others<sup>a</sup>, n (%) | 6,694 (6.4) | 6,158 (6.3) | 536 (7.2) |
| TDI, mean ± SD | -1.8±2.8 | -1.8±2.8 | -1.7±2.9 |
| BMI, kg/m<sup>2</sup>, mean ± SD | 26.7±4.5 | 26.6±4.5 | 28.3±4.7 |
| Smoking status |  |  |  |
| Non-smoker, n (%) | 61,192 (58.3) | 57,759 (59.2) | 3,433 (46.4) |
| Ever smoker, n (%) | 36,383 (34.7) | 33,012 (33.8) | 3,371 (45.6) |
| Current smoker, n (%) | 7,364 (7.0) | 6,770 (6.9) | 594 (8.0) |
| Alcohol consumption |  |  |  |
| Never or occasional, n (%) | 14,999 (14.3) | 13717 (14.1) | 1282 (17.3) |
| 1-3 time/month, n (%) | 11,381 (10.8) | 10641 (10.9) | 740 (10.0) |
| 1-4 times/week, n (%) | 53,848 (51.3) | 50312 (51.6) | 3536 (47.8) |
| Daily, n (%) | 24,711 (23.5) | 22871 (23.4) | 1840 (24.9) |
| Regular exercise, yes, n (%) | 60,010 (57.2) | 55965 (57.4) | 4045 (54.7) |
| Maternal smoking around birth, yes, n (%) | 30,191 (28.8) | 27805 (28.5) | 2386 (32.3) |
| Family history of CVD, yes, n (%) | 60,254 (57.4) | 54922 (56.3) | 5332 (72.1) |

Notes: PhenoAge, Phenotypic Age; TDI, Townsend deprivation index; BMI, body mass index; CVD, cardiovascular disease.
<sup>a</sup>Others included unemployed, student, volunteer, and so on.

### Associations of childhood, adulthood, and lifetime traumas with CVD

The Shapley Value Decomposition method suggested that childhood traumas, adulthood traumas, and lifetime traumas contributed to 1.0%, 0.9%, and 15.2% of differences in CVD prevalence, respectively (**Figure S1**). **Table 2** shows the associations of traumas in childhood, adulthood, and lifetime with CVD. We found that when compared with subgroups who experienced mild childhood traumas, those who experienced severe childhood traumas had a higher odd of CVD, with an aOR of 1.38 (95% CI: 1.30, 1.46) after adjustment for chronological age and sex (Model 1). Also, for adulthood traumas, lifetime traumas, and all traumas, compared to those who experienced mild traumas, participants who experienced severe traumas had 22%, 102%, and 55% increased odds of CVD, respectively (Model 1). After additionally adjusting for ethnicity, educational level, occupation, TDI, BMI, smoking status, alcohol consumption, regular exercise, maternal smoking around birth, and family history of CVD, the strength of these associations slightly decreased (2%-20%) but remained statistically significant (Model 2). These associations persisted after further adjustment for PhenoAge (Model 3).

**Table 2.**
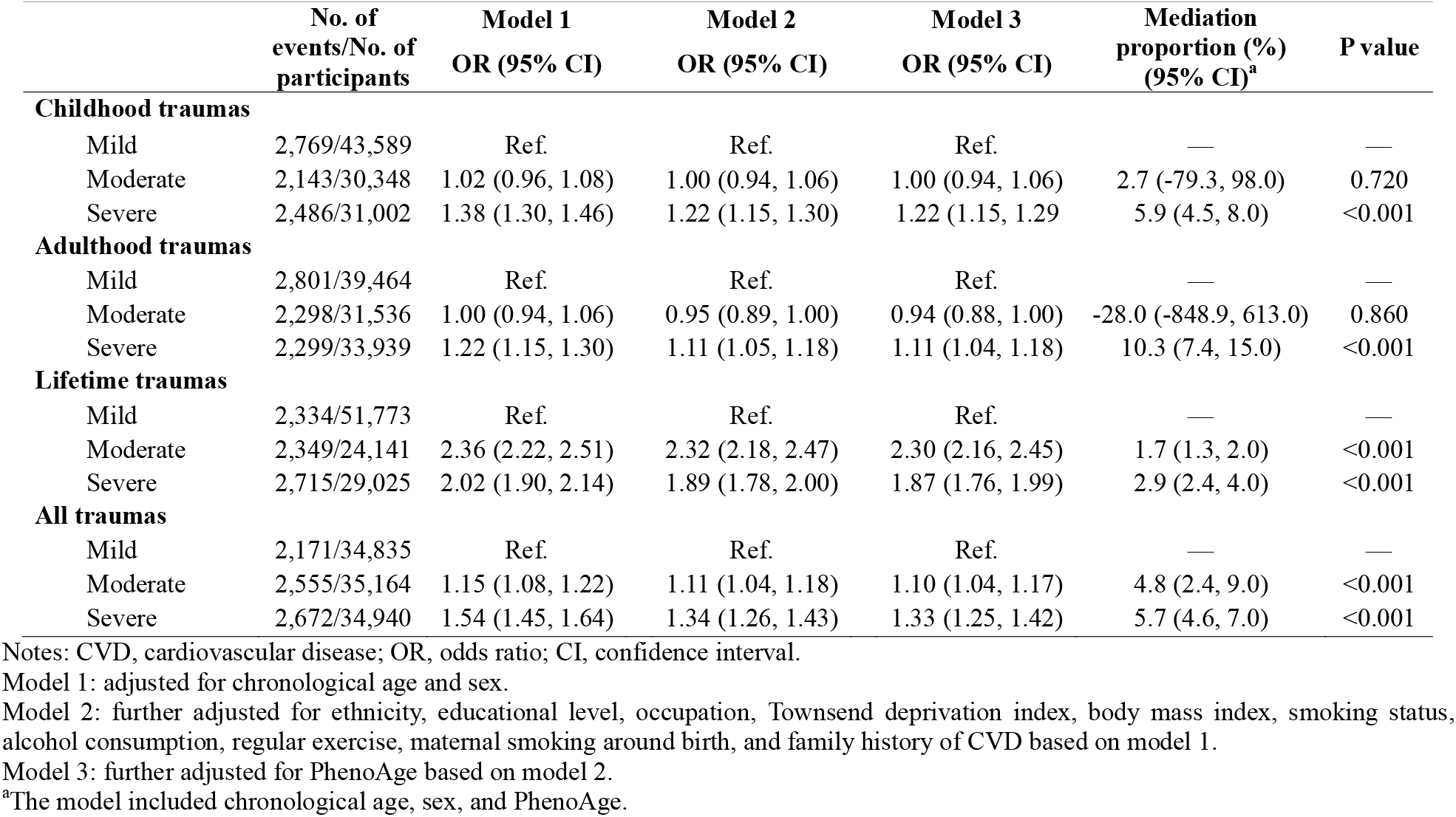
Associations of life course traumas with CVD and mediation proportion of life course traumas in CVD attributed to phenotypic aging.

### Associations of life course traumatic profiles with CVD

**Table 3** shows the significant associations of life course traumatic profiles with CVD. Compared with subgroups experiencing *non-severe traumas across life course*, those experiencing *non-severe childhood and severe adulthood traumas, severe childhood and non-severe adulthood traumas*, and *severe traumas across life course* had higher odd of CVD, with aORs of 1.07 (95% CI: 1.00, 1.15), 1.17 (95% CI: 1.09, 1.25), and 1.33 (95% CI: 1.24, 1.43), respectively (Model 3).

**Table 3.** Associations of four life course traumatic profiles with CVD and mediation proportion of life course traumatic profiles in CVD attributed to phenotypic aging.

|  | No. of events/No. of participants | Model 1<br>OR (95% CI) | Model 2<br>OR (95% CI) | Model 3<br>OR (95% CI) | Mediation proportion<br>(%) (95% CI)<sup>a</sup> | P<br>value |
| --- | --- | --- | --- | --- | --- | --- |
| *Non-severe traumas across life course* | 3,788/55,094 | Ref. | Ref. | Ref. | — | — |
| *Non-severe childhood and severe adulthood traumas* | 1,124/18,843 | 1.11 (1.03, 1.19) | 1.07 (1.00, 1.15) | 1.07 (1.00, 1.15) | 6.3 (2.2, 22.0) | 0.008 |
| *Severe childhood and non-severe adulthood traumas* | 1,311/15,906 | 1.29 (1.20, 1.38) | 1.17 (1.09, 1.25) | 1.17 (1.09, 1.25) | 4.6 (2.9, 7.0) | <0.001 |
| *Severe traumas across life course* | 1,175/15,906 | 1.56 (1.45, 1.67) | 1.34 (1.24, 1.44) | 1.33 (1.24, 1.43) | 6.4 (5.2, 8.0) | <0.001 |

Notes: CVD, cardiovascular disease; OR, odds ratio; CI, confidence interval.
Model 1: adjusted for chronological age and sex.
Model 2: further adjusted for ethnicity, educational level, occupation, Townsend deprivation index, body mass index, smoking status, alcohol consumption, regular exercise, maternal smoking around birth, and family history of CVD based on model 1.
Model 3: further adjusted for PhenoAge based on model 2.
<sup>a</sup>The model included chronological age, sex, and PhenoAge.

### Mediation analyses of phenotypic aging on the associations of life course traumas with CVD

First, the associations of phenotypic aging with CVD were presented in **Table S4**. With adjustment for chronological age and sex, a 1-year increase in PhenoAge increased the odd of CVD by 5% (P<0.001). The result remained unchanged after further adjustment for other covariates. Additionally, the associations of PhenoAge with CVD stratified by subgroups of childhood traumas, adulthood traumas, lifetime traumas, and all traumas were estimated (**Figure 3**). We did not observe remarkable changes in the results. For example, when stratifying participants by subgroups of lifetime traumas, PhenoAge was consistently associated with CVD after adjustment for chronological age, sex, ethnicity, educational level, occupation, TDI, BMI, smoking status, drinking status, regular exercise, maternal smoking around birth, and family history of CVD, with aORs of 1.02 (95% CI: 1.01, 1.03), 1.03 (95% CI: 1.02, 1.04), and 1.03 (95% CI: 1.02, 1.04) in mild, moderate and severe subgroups, respectively.

**Figure 3.**
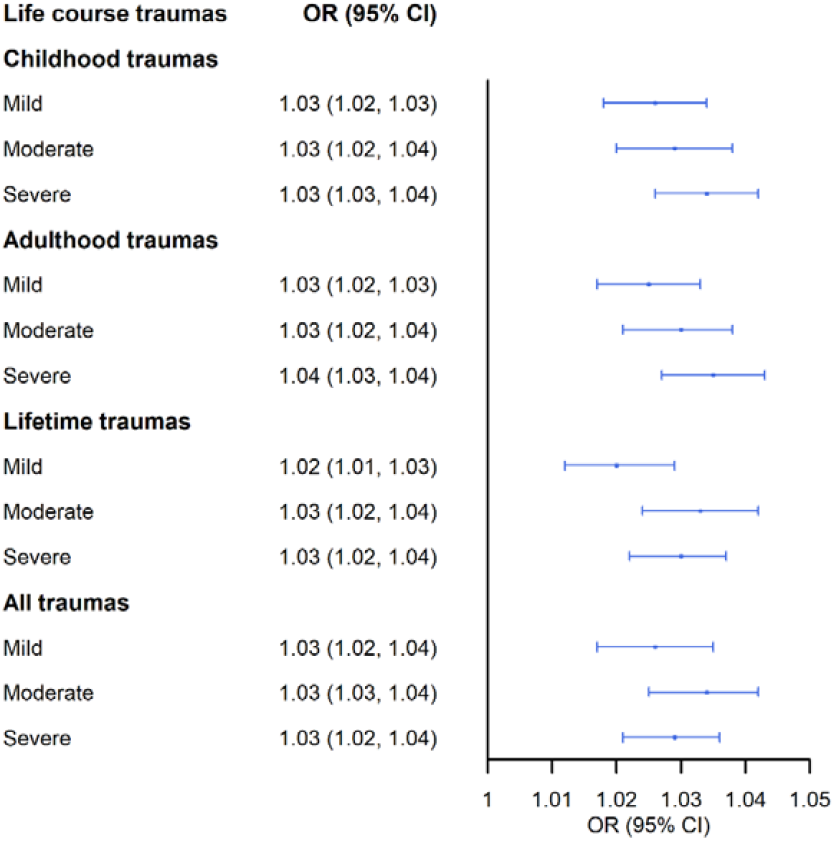
Associations of phenotypic aging with CVD by life course traumas subgroups. Odds ratios were adjusted for chronological age, sex, ethnicity, educational level, occupation, Townsend deprivation index, body mass index, smoking status, drinking status, regular exercise, maternal smoking around birth, and family history of CVD. OR, odds ratio; CI, confidence interval; CVD, cardiovascular disease.

Next, as shown in **Table S5**, compared with those who experienced mild childhood traumas, subgroups who experienced severe childhood traumas were significantly associated with PhenoAge, with a β of 0.06 (standard error [SE]=0.04, P=0.089) (Model 2). The corresponding values for adulthood traumas, lifetime traumas, and all traumas were 0.16 (SE=0.04, P<0.001), 0.19 (SE=0.04, P<0.001), and 0.11 (SE=0.04, P<0.001), respectively (Model 2).

Finally, **Table 2** presents the results of formal mediation analyses for childhood traumas, adulthood traumas, lifetime traumas, and all traumas. Overall, the results suggested that when comparing those in mild subgroup for childhood traumas, adulthood traumas, lifetime traumas, and all traumas to those in the severe subgroup, 5.9% (95% CI: 4.5%, 8.0%; P<0.001), 10.3% (95% CI: 7.4%, 15.0%; P<0.001), 2.9% (95% CI: 2.4%, 4.0%; P<0.001), and 5.7% (95% CI: 4.6%, 7.0%; P<0.001) of CVD events were mediated by PhenoAge, respectively. Also, PhenoAge mediated 6.4% (95 CI: 2.2%, 22.0%; P=0.008), 4.6% (95% CI: 2.9%, 7.0%; P<0.001) and 6.4% (95% CI: 5.2%, 8.0%) of increased CVD events in those who experienced *non-severe childhood and severe adulthood traumas, severe childhood and non-severe adulthood traumas, and severe traumas across life course*, relative to subgroups experiencing *non-severe traumas across life course* (**Table 3**).

### Sensitivity analyses

First, differences in socio-demographic characteristics between included participants and those excluded from this study in UKB were found (**Table S6**). Participants who were included were more likely to be younger, female, White, non-smoker, and daily drinker, to have a higher percentage of high educational level, working, and regular exercise, a lower percentage of maternal smoking around birth, and lower TDI and BMI. Second, we estimated the associations of the single traumas item and summary score of all traumas with CVD (**Table S7**). Almost all traumas items were significantly associated with CVD, except for the three lifetime traumas items (i.e., Been involved in combat or exposed to war-zone, victim of physically violent crime, and witnessed sudden violent death) that were uncommon. Third, associations of nine profiles of life course traumas with CVD are presented in **Table S8**. We found that those who experienced severe childhood traumas had persistently increased odds of CVD. Compared with *Mild-Mild* profile, the odds of CVD for *Severe-Mild, Severe-Moderate*, and *Severe-Severe* profiles were 1.13 (95% CI: 1.02, 1.26), 1.10 (95% CI: 1.00, 1.21), and 1.28 (95% CI: 1.17, 1.39), respectively (Model 2). PhenoAge mediated 3.1% (95% CI: 2.2%, 7.0%; P=0.032), 10.7% (95% CI: 6.8%, 18.0%; P<0.001), and 8.0% (95% CI: 6.3%, 10.0%; P<0.001) of increased CVD events in those who experienced *Severe-Mild, Sever-Moderate*, and *Severe-Severe* traumatic profiles, relative to those who experienced *Mild-Mild* traumatic profile. These results were consistent with those based on four life course traumatic profiles. Additionally, the *Severe-Severe* profile was substantially associated with PhenoAge, with a β of 0.19 (SE=0.05, P<0.001) (**Table S9**). Fourth, the exclusion of 996 self-reported CVD did not substantially change the significant associations of life course traumas and CVD (**Table S10**). For example, compared with subgroups who experienced mild lifetime traumas, those experiencing moderate and severe lifetime traumas had increased odds of CVD, with aORs of 2.49 (95% CI: 2.33, 2.66), and 1.89 (95% CI: 1.77, 2.02), respectively (**Table S10**, model 3). Fifth, we estimated the associations of childhood traumas and incident CVD that ascertained after baseline. The results did not change substantially. Compared with subgroups who experienced mild childhood traumas, those experiencing severe childhood traumas had an increased risk of CVD, with an adjusted hazard ratio of 1.18 (95% CI: 1.11, 1.25) (**Table S11**). Finally, we found similar results when categorizing childhood, adulthood, lifetime, and all traumas into three subgroups according to the tertile of original summary score, respectively (**Table S12** and **Table S13**). For instance, PhenoAge mediated 17.1%, 3.5%, and 7.9% of increased CVD events in subgroups who experienced *non-severe childhood and severe adulthood traumas, severe childhood and non-severe adulthood traumas*, and *severe traumas across life course*, relative to those experiencing *non-severe traumas across life course*, respectively (**Table S13**).

## Discussion

Based on a large sample of adults from the UK, we found that childhood, adulthood, and lifetime traumas, as well as diverse life course traumatic profiles, were significantly associated with CVD. Furthermore, using a well-developed phenotypic aging measure — PhenoAge, we demonstrated that phenotypic aging partially mediated these associations, providing evidence for underlying pathways that link life course trauma to CVD. These findings highlight the importance of preventive programs targeting traumas throughout the life course, aiming to slow phenotypic aging and further promoting cardiovascular health.

The findings on childhood traumas are consistent with those from previous studies[2-5]. The greatest contribution of this study was that we incorporated traumas over the life course, allowing us to estimate the influence of both childhood and adulthood traumas on cardiovascular health inequalities in the same sample. The multivariable logistic models indicated that childhood and adulthood traumas are all significantly associated with CVD. In comparison to the traditional method (e.g., multivariable logistic models), the Shapley Value Decomposition method estimated relatively accurate contributions of childhood and adulthood traumas simultaneously. In this study, we estimated that inequalities in cardiovascular health contributed by traumas in childhood was 0.1% higher than that in adulthood. The associations of childhood traumas and adulthood traumas with CVD were further confirmed. Moreover, the findings that *severe childhood and non-severe childhood traumas*, and *severe traumas across life course* profiles have relatively higher odds of CVD with comparison to *non-severe traumas across life course* profile, suggesting that long-lasting negative influence of severe childhood traumas cannot be ameliorated even when adults experienced non-severe traumas in adulthood. Life course approach enhances our perception to inform preventive and intervention programs at the window phase that diminish the onset and development of CVD for a lifetime. The chains of life course exposures emphasize not only the underlying process of the accumulative risk model, but also specific patterns of the critical periods model [29]. Thus, government policies and preventive programs should target traumatic events over the life course, particularly in childhood, to reduce CVD events.

Lifetime traumas in our study tracked the accumulative effect of traumas from the perspective of a life course theory. This may explain why lifetime traumas contributed most to CVD (15.2%). Nevertheless, we must acknowledge the existence of selection bias [30, 31], especially for the survivor bias, which means that it was hard to include participants who were exposed to the most severe traumatic experiences (i.e., more severe than severe subgroup in our study) in early life. In our study, few participants reported a high frequency of traumatic experiences. Despite the existing limitation, our results consistently suggest that childhood, adulthood, and lifetime traumas have their influences on CVD, affirming the great value of a life course approach.

In general, we verified our proposed hypotheses: life course traumas influence CVD through different rates of aging. Furthermore, the formal mediation analyses provided further evidence for the “get under the skin” hypothesis. The potential mechanism suggests that slowing phenotypic aging would probably help to narrow the gap of cardiovascular health inequalities, which is in line with the Geroscience [11, 32]. The development of aging measures is still in rapid progress, particularly using molecular markers (e.g., DNA methylation), and thus will provide us with better tools to comprehensively understand the role of aging in the development of disease. Despite the unavailability of such data in UKB, we call for more studies to use various aging measures to validate our observations in this study.

Our findings for childhood, adulthood, and lifetime traumas highlight the importance of preventing traumas over the life course in reducing CVD events, while the findings for diverse life course traumatic profiles implicate us that traumatic events that occurred at early stage may have long-lasting influence on CVD. The family and social community should keep a watchful eye on the occurrence of traumatic events beginning from the childhood period. Furthermore, findings from the mediation analyses suggest that geroprotective programs may also help prevent CVD, despite to a lesser extent. Researchers have been making efforts toward this direction, such as studying the protective effect of anti-aging drugs (e.g., chondroitin sulfate, metformin, rapamycin, and resveratrol) [33-36], which is promising.

Major strengths of this study included the large sample size from a national survey of UK middle-aged and older participants, and a unique dataset about traumas over the life course. Furthermore, formal mediation analyses were performed to assess the pathway from life course traumas to CVD through phenotypic aging. However, several limitations should be noted. First, information on life course traumas were obtained by self-report in the year 2016, which may result in recall bias. But recent studies have shown that similar information about the stress-buffering process was provided using retrospective and prospective methods [37, 38]. Second, participants in this study were not representative of general adults in the UK. The majority of participants were White, had higher socio-economical levels, and tended to be healthier (i.e., “healthy volunteer” effect) [30], leading to possible underestimation of these associations. These results may not be generalizable to other populations, such as populations from developing countries. Fourth, due to the short-term follow-up after the 2016 online mental health questionnaire survey, we did not estimate the influence of life course traumas and PhenoAge on the incidence of CVD. Fifth, information on frequencies, instead of severity, of life course traumas were collected. Finally, this was a retrospective study design based on the existing UKB data. In moving forward, prospective study design should be considered to reinforce our findings.

In this large sample of middle-aged and older adults from the UK, childhood, adulthood, and lifetime traumas, as well as diverse life course traumatic profiles, were associated with CVD. This study also showed that phenotypic aging partially mediated the associations between life course traumas and CVD, providing evidence to improve our understanding of the pathway from life course traumas to CVD. These findings underscore the importance of policy programs targeting traumatic events over the life course in ameliorating inequalities in aging and cardiovascular health.

## Data Availability

Data from UK Biobank are available on application at www.ukbiobank.ac.uk/register-apply.

## Author contributions

ZL conceived and designed the study. XC, JZ, and CM performed the analysis. XC wrote the initial draft of the manuscript. XL, CK, ML, GH, HA, Xi Chen, XW, and ZL helped to interpret the results and edit the manuscript. CK, ML, GH, HA, Xi Chen, XW, and ZL contributed to the critical revision of the manuscript for important intellectual contents. All authors reviewed and approved the final version of the manuscript.

## Data availability statement

Data from UK Biobank are available on application at www.ukbiobank.ac.uk/register-apply.

## Acknowledgments

This research has been conducted using the UK Biobank resource under application number 61856. We wish to acknowledge the UK Biobank participants who provided the sample that made the data available.

